# TONSILS ARE MAJOR SITES OF PROLONGED SARS-COV-2 INFECTION IN CHILDREN

**DOI:** 10.1101/2023.01.21.23284592

**Authors:** Thais M. Lima, Ronaldo B. Martins, Carolina S. Miura, Maria V. O. Souza, Murilo H. A. Cassiano, Tamara S. Rodrigues, Flávio P. Veras, Josane F. Sousa, Rogério Gomes, Glaucia M. Almeida, Stella R. Melo, Gabriela C. Silva, Matheus Dias, Carlos F. Capato, Maria L. Silva, Veridiana E. D. Barros, Lucas R. Carenzi, Dario S. Zamboni, Daniel M. M. Jorge, Edwin Tamashiro, Wilma T. Anselmo-Lima, Fabiana C. P. Valera, Eurico Arruda

**Affiliations:** Department of Cell Biology and Virology Research Center, University of Sao Paulo School of Medicine, Ribeirão Preto, Brazil; Department of Clinical Analyses, Toxicology and Food Science, University of Sao Paulo, School of Pharmaceutical Science of Ribeirão Preto, Brazil; Department of Ophthalmology, Otorhinolaryngology and Head and Neck Surgery, University of Sao Paulo School of Medicine, Ribeirão Preto, Brazil

**Author notes:** Corresponding author: Eurico Arruda; Department of Cell Biology, University of São Paulo School of Medicine, Av. Bandeirantes 3900, Ribeirão Preto, SP, Brazil, 14049-900. These authors contributed equally to this work (Thais M. Lima, Ronaldo B. Martins, Carolina S. Miura, Fabiana C. P. Valera, Eurico Arruda).

## Abstract

In the present study, we show that SARS-CoV-2 can infect palatine tonsils and adenoids in children without symptoms of COVID-19, with no history of recent upper airway infection. We studied 48 children undergoing tonsillectomy due to snoring/OSA or recurrent tonsillitis between October 2020 and September 2021. Briefly, nasal cytobrush (NC), nasal wash (NW) and tonsillar tissue fragments obtained at surgery were tested by RT-PCR, immunohistochemistry (IHC), flow cytometry and neutralization assay. We detected the presence of SARS-CoV-2 in at least one specimen tested in 25% of patients (20% in palatine tonsils and 16.27% in adenoids, 10.41% of NC and 6.25% of NW). Importantly, in 2 of the children there was evidence of laboratory-confirmed acute infection 2 and 5 months before surgery. IHC revealed the presence of SARS-CoV-2 nucleoprotein in epithelial surface and in lymphoid cells in both extrafollicular and follicular regions, in adenoids and palatine tonsils. Flow cytometry showed that CD20^+^ B lymphocytes were the most infected phenotypes by SARS-CoV-2 NP, followed by CD4+ and CD8+ T lymphocytes, and CD14+ macrophages and dendritic cells. Additionally, IF indicated that SARS-CoV-2-infected tonsillar tissues had increased expression of ACE2 and TMPRSS2. NGS sequencing demonstrated the presence of different SARS CoV-2 variants in tonsils from different tissues. SARS-CoV-2 antigen detection was not restricted to tonsils, but was also detected in nasal cells from the olfactory region. In conclusion, palatine tonsils and adenoids are sites of prolonged infection by SARS-CoV-2 in children, even without COVID-19 symptoms.

## 1. INTRODUCTION

SARS-CoV-2 causes COVID-19, a respiratory disease affecting people of all ages worldwide.^1^ However, severe COVID-19 is much less frequent in children and adolescents than in adults.^2^ The reasons for children having less severe disease are not entirely understood, but lower expressions of ACE2 and TMPRSS2 in children’s respiratory tract are contributing factors.^3^

We have previously reported high rates of PCR detection of respiratory viruses, including endemic coronaviruses, in tonsils and adenoids from patients with chronic tonsillar diseases, without recent history of symptomatic airway infections.^4^ This finding was later confirmed by other groups,^5,6,7^ indicating that human tonsils are sites of asymptomatic respiratory virus infections. This prompted us to test whether SARS-CoV-2 was also detected in children’s tonsils during the COVID-19 pandemic. Here we report detection of SARS-CoV-2 RNA and protein in tonsils, nasal cytobrushes, and respiratory secretions from children with tonsillar hypertrophy lacking symptoms of COVID-19, and determine the types of infected cells.

## 2. PATIENTS AND METHODS

### 2.1. Study design and sample processing

This cross-sectional study was done from October 2020 to September 2021, at the Otorhinolaryngology Division of the School of Medicine of Ribeirão Preto, University of São Paulo, and enrolled 3 to 11-year-old children who underwent adenoidectomy and/or tonsillectomy to treat recurrent tonsillitis or obstructive sleep apnea. The study was approved by the Ethics Committee of the Clinical Hospital, Ribeirao Preto Medical School, University of São Paulo. Exclusion criteria were symptoms of acute respiratory infections in the month before surgery, craniofacial malformations, genetic syndromes, deposit diseases, immunodeficiencies, and suspected tonsillar cancer. None of the children had been vaccinated, since COVID-19 vaccines had not been approved in Brazil at that time.

On the day of surgery parents/guardians who agreed to participate signed informed consents. A questionnaire was filled with data on previous exposure to COVID-19, demographics, comorbidities, indications for tonsillectomy, and relevant physical examination findings, and recorded using REDCap software (Vanderbilt University, USA).

During surgery, the following samples were obtained: bilateral nasal wash (10 ml of saline solution instilled and immediately aspirated); bilateral cytobrush of the olfactory area under a 0º rigid optical view (a small brush was positioned close to the olfactory fossa, rotated ten times, and immediately placed into sterile RPMI medium with 4% antibiotic/antimycotic solution (Gibco, Gran Island, USA); 4 ml of peripheral blood for serology; adenoid and palatine tonsil tissues immediately placed in the same medium used for the cytobrushes. All samples were transported to the laboratory on ice within 2 hours.

The tissue specimens were washed in PBS to remove debris and blood clots, and used to prepare four aliquots for: qRT-PCR, virus isolation in cell culture, purification of tonsillar mononuclear cells (TMNC), and fixation for histological tests. Tissue pieces of ∼0.25□cm^3^ were treated with collagenase type I (100 U/ml) and dispase (0.6 U/ml) for 1h at 37°C (Gibco, Grand Island, USA), then passed through a nylon mesh to obtain a cell suspension, which was used for: 1) mixing with 2X viral transport medium (VTM), consisting of minimum essential medium (MEM) with 20% FBS and 15% glycerol for viral isolation; 2) RNA extraction using Trizol; 3) backup in RNA-later (Invitrogen); 4) isolation of TMNCs by Ficoll-Paque™, which were suspended in freezing medium (RPMI with 20% FBS and 10% DMSO). Aliquots were stored at −80°C. Another piece of tonsillar tissue was placed in Carnoy’s fixative for 12h and embedded in paraffin. The nasal washes and cytobrushes were used to prepare aliquots in VTM and Trizol, and the remainder was used to prepare cell pellets that were spotted onto glass slides, fixed with acetone and stored at -20°C for immunofluorescence.

### 2.2. Quantitation of SARS-CoV-2 RNA

SARS-CoV-2 RNA was quantitated by real-time RT-PCR (qRT-PCR) with primers and probes for the N2 and E genes, according to protocols proposed by the USA Centers for Disease Control and Prevention, and the Charité Group (Table 1). The one-step reaction was assembled with 1000 ng of total RNA, specific primers (20 μM) and probe (5 μM), and the qPCRBIO Probe 1-Step Go-PCR Biosystems (Applied Biosystems, Foster City, USA) with the parameters: 55°C for 10 min, 95°C for 2 min, 40 cycles of 95°C for 5□s, 60°C for 30□s, on a Step-One Plus (Applied Biosystems).

**Table 1.**
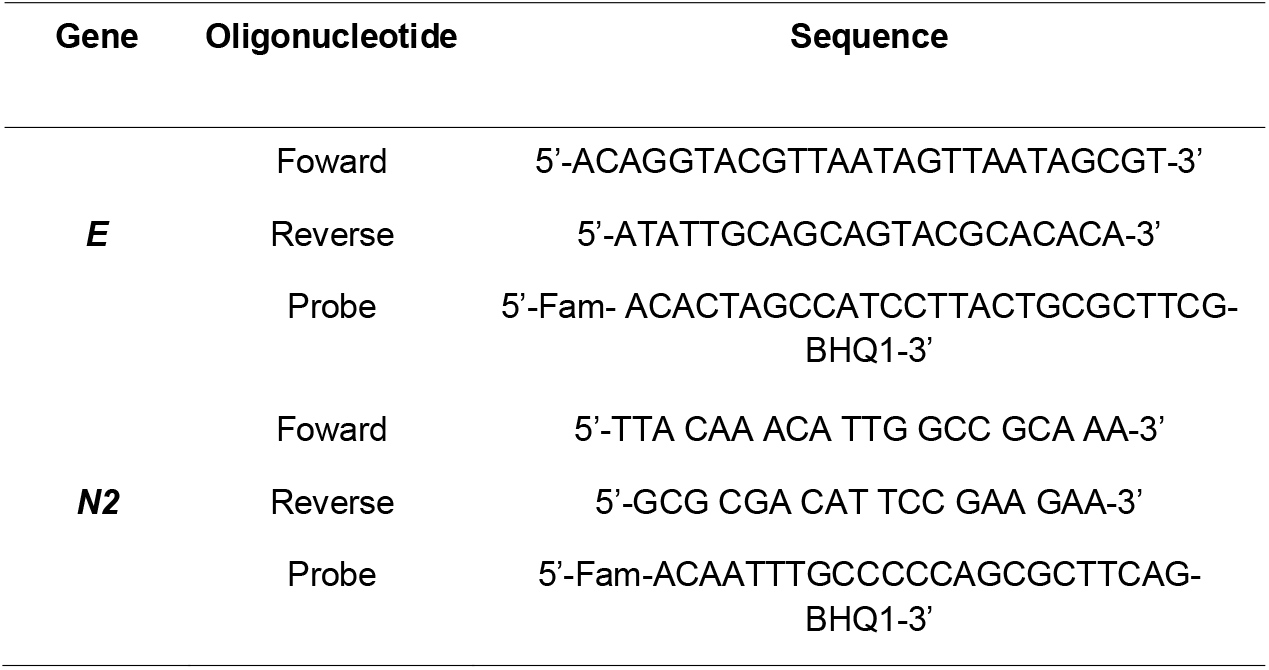
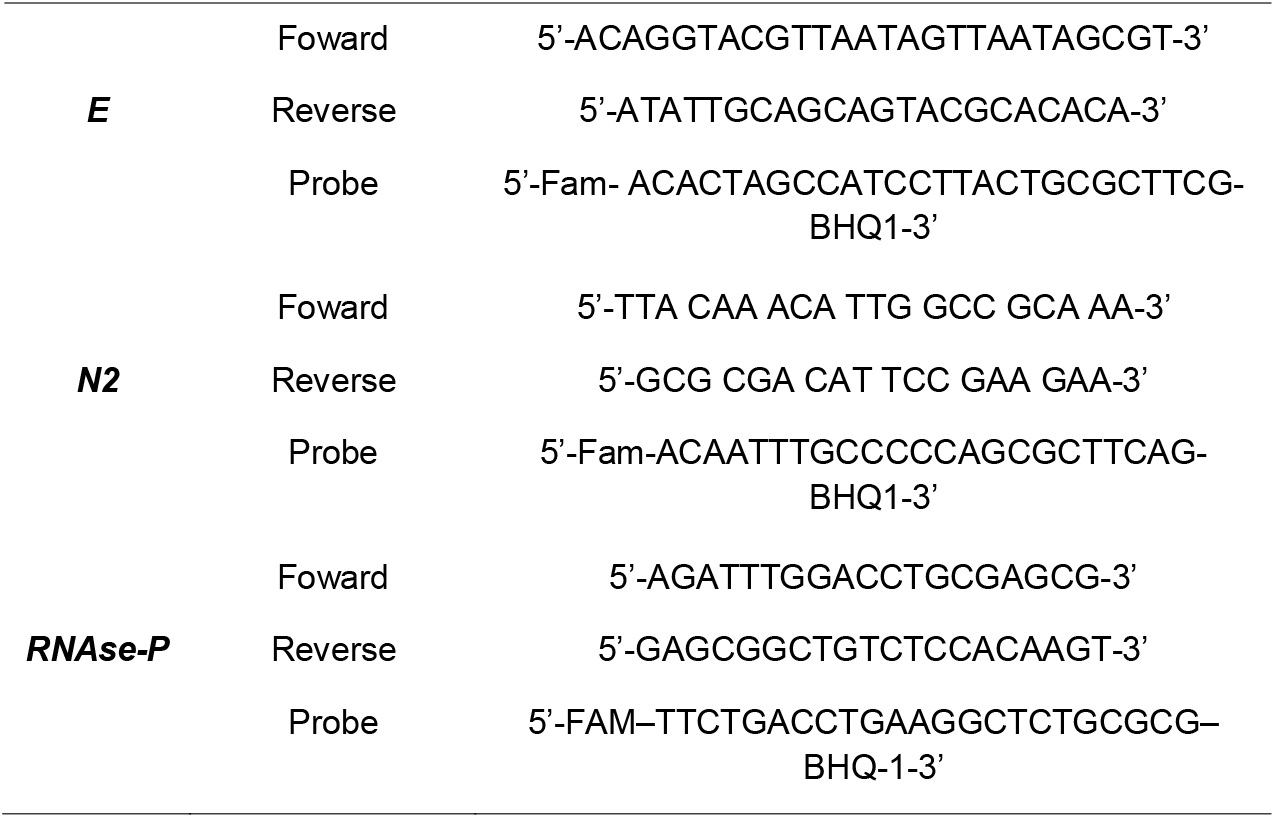
Sets of primers and probes for the detection of SARS-CoV-2 by RT-PCR.

SARS-CoV-2 qRT-PCR was done in triplicate and results were normalized by amplification of the RNAseP gene. SARS-CoV-2 genome loads were extrapolated using a standard curve prepared with a plasmid containing a 944 bp amplicon of the N gene, inserted in the TA cloning vector (PTZ57R/T CloneJetTM Cloning Kit (Thermo Fisher®). Viral RNA loads were plotted with GraphPad® Prism 8.4.2 194 software.

### 2.3. Immunohistochemistry for SARS-CoV-2 antigen in adenotonsillar tissue

SARS-CoV-2 antigen was detected *in situ* in tissue sections by immunohistochemistry (IHC). IHC standardization was done with Vero CCL-81 cells infected with parental SARS-CoV-2 Wuhan lineage as positive controls. Tissue sections (3μm) were deparaffinized, rehydrated and subjected to the previously published protocols.^8^ Red-purple signal was developed with AEC peroxidase system kit (SK-4800, Vector Laboratories, Burlingame, CA) and sections were counterstained with Harris hematoxylin (Vector) and scanned with a ScanScope VS120 (Olympus Life Sciences, Tokyo, Japan) in bright field, using 400× magnification.

### 2.4. Immunofluorescence for ACE2, TMPRSS2 and SARS-CoV-2 S protein in tonsils

Tissue sections were deparaffinized in three xylene baths (Synth, Diadema, Brazil) for 5 minutes each, and hydrated by sequential incubations of 3 minutes each in decreasing ethanol concentrations (JTBacker) (100%, 90%, 80%, 70%, and 50%). Sections were blocked for 20 minutes in PBS with 0.01% BSA (Gibco) before antibody incubation. Slides were stained for 1 hour with the following primary antibodies diluted in PBS-BSA: rabbit monoclonal anti-SARS-CoV-2 S protein (Invitrogen) diluted 1:500; goat polyclonal anti-ACE2 (R&D systems) diluted 1:200; or mouse monoclonal anti-TMPRSS2 (Millipore) diluted 1:200. After washing with Tris-buffered saline with 0.1% Tween-20 (TBST), slides were incubated with AlexaFluor 488-labeled secondary alpaca anti-mouse IgG antibodies (Jackson ImmunoReseacher) diluted 1:1000, or alpaca anti-rabbit IgG AlexaFluor 594 (Jackson ImmunoReseacher) diluted 1:1000. Nuclei were stained with DAPI (Vector) and images were acquired by Axio Observer combined with LSM 780 confocal microscope (Carl Zeiss) at 63x magnification at the same setup of zoomed and laser rate. Ten random fields per sample were analyzed at the x and y focal planes to measure the mean fluorescence intensity of ACE2 and TMPRSS2. Images were analyzed using Fiji by Image J.

### 2.5. Indirect immunofluorescence staining for SARS-CoV-2 N protein in nasal cytobrush samples

Slides were defrosted and incubated with a permeabilizing/blocking solution of PBS with 0.01% Triton, 1% BSA (Sigma) and 5% goat serum for 5 min at room temperature. Slides were washed in PBS and incubated with primary rabbit polyclonal antibody anti-SARS-CoV-2 N protein (Creative Diagnostics) at 37°C for 1 hour. After blocking with SuperBlock™, slides were incubated with a Alexa 594-conjugated goat anti-rabbit secondary antibody (Abcam). The nuclei were stained with DAPI (Thermo Fisher), and images were obtained with a confocal fluorescence Leica TCS SP8 microscope (Leica Microsystems).

### 2.6. Flow cytometry of dissociated tonsillar lymphomononuclear cells

Frozen purified TMNCs were analyzed by flow cytometry after a 30 min staining at 4°C using antibodies for CD4 (PerCP-Cy5.5), CD8 (PE-Cy7), CD11c (PE-Cy7), CD14 (PerCP), CD20 (PE-Cy7) and CD123 (PerCP-Cy5.5) (BD Pharmingen). Cells were then washed, permeabilized and fixed with BD Cytofix/Cytoperm™ kit.

Intracellular SARS-CoV-2 was stained with rabbit anti-SARS-CoV-2 NP antibody for 1□h, followed by anti-rabbit IgG-APC secondary antibody (BD Pharmingen) for 30 min. TMNCs obtained from tonsils RT-PCR-negative for SARS-CoV-2 were used as negative controls. Cell preparations stained only with the IgG-APC secondary antibody were used for calibration of PE acquisition. Acquisition was performed in fixed cells in a flow cytometer (BD Accuri C6; BD Biosciences) and then analyzed using FlowJo software (Tree Star).

### 2.7. Serology assays

COVID-19 IgM/IgG detections in patients sera were done using two rapid test kits from Nantong Egens Biotechnology and Genrui Biotech Inc, following the manufacturer’s instructions. Determination of 100% virus neutralization titer (VNT_100_) was done by the same protocol we previously published.^9^

### 2.8. Sequencing of SARS-CoV-2 genomes

SARS-CoV-2 genome sequencing followed the ARTIC nCoV-2019 protocol v3 (https://protocols.io/view/ncov-2019-sequencing-protocol-v3-locost-bh42j8ye)^10^ with PCR cycling modifications previously proposed.^11^ Briefly, reverse transcription was performed with the LunaScript RT SuperMix Kit (NEB), using total RNA from tonsillar tissues and cytobrushes that had yielded Ct values ≤38 for genes E and N by qRT-PCR. Two multiplexed PCR products (Pool A = 110 primers pairs and Pool B = 108 primers pairs) were generated using the Q5 Hot Start High-Fidelity DNA Polymerase (NEB) and the primer scheme from IDT ARTIC nCoV-2019 V3 Panel. PCR products were purified using Agencourt AMPure XP beads (Beckman Coulter™) and the DNA concentration was measured by the Qubit 2 Fluorometer using the Qubit dsDNA HS Assay Kit (Invitrogen). DNA products (Multiplex PCR pools A and B) were pooled together in a final concentration of 50 fmol. The Ligation Sequencing kit used in the MinION library preparation was the SQK-LSK-109 and Native Barcoding was done with the NANO-EXPNBD196 kit (Oxford Nanopore, Oxford, UK). The obtained library was loaded on R9.4 Oxford MinION flowcells (FLO-MIN106) and sequenced using the MinION Mk1B device.

### 2.10. Bioinformatic analysis and sequence availability

The pipeline included (i) the ONT MinKNOW software for collection of raw data and quality control, and (ii) Guppy (v6.0.1) for high accuracy base calling. Assembly of the high-accuracy base called Fastq files was done by the nCoV-2019 novel coronavirus bioinformatics protocol (https://artic.network/ncov-2019/ncov2019-bioinformatics-sop.html accessed on January 12, 2022) with Minimap2^12^ and Rampart^13^ for genome coverage analysis.

The assembled genomes were analyzed with Nextclade v1.14.0 (https://clades.nextstrain.org/ accessed on August 26, 2022)^14^ and Pangolin v4.0 (https://github.com/cov-lineages/pangolin accessed on January 20, 2022)^15^ to identify the clade and lineages.

Raw Minion Nanopore reads from experiment were submitted to the SRA NCBI database under the Bioproject ID PRJNA876260, and the following BioSample IDs: SAMN30649069, SAMN30649070, SAMN30649072, SAMN30649075, SAMN30649076, SAMN30649077, SAMN30649078, SAMN30649079, SAMN30649080, and SAMN30649081. Controll raw Minion Nanopore reads were submitted to the SRA NCBI database under the Bioproject ID PRJNA909758, and the following BioSample IDs: SAMN32093249.

### 2.11. Statistical analysis

Continuous data was analyzed by Student t test or Mann–Whitney test, depending on normal distribution. Statistical tests and graph plotting were performed with GraphPad Prism 8.4.2 software.

## 3. RESULTS

### 3.1. Demographic characteristics

A total of 48 patients (caucasian: 57.2%) were enrolled in the study. The patients were aged 3 to 11 years (mean 5.9 ± 2.2); 30 were boys (62.5%), and 24 (50%) did not have associated diseases (Table 2). Among the reported comorbidities, allergic rhinitis was reported in 19 children (39.6%), recurrent otitis media in 6 (12.5%), and mild asthma in 4 (8.3%). According to the parents/guardians, the last acute upper airways infection requiring or not antibiotics occurred 1 to 24 (average 9.2) months before surgery. Eight children (17%) had been exposed to confirmed COVID-19 in the household forty days to six months before surgery. Two patients had previous laboratory-confirmed SARS-CoV-2 infection: one had a positive IgM test three months before surgery, and the other had a positive RT-PCR in respiratory secretion five months before surgery. An additional child reported smell and taste changes that occurred more than one month prior surgery, but was not tested for SARS-CoV-2 (Supplementary Table 1).

**Table 2.**
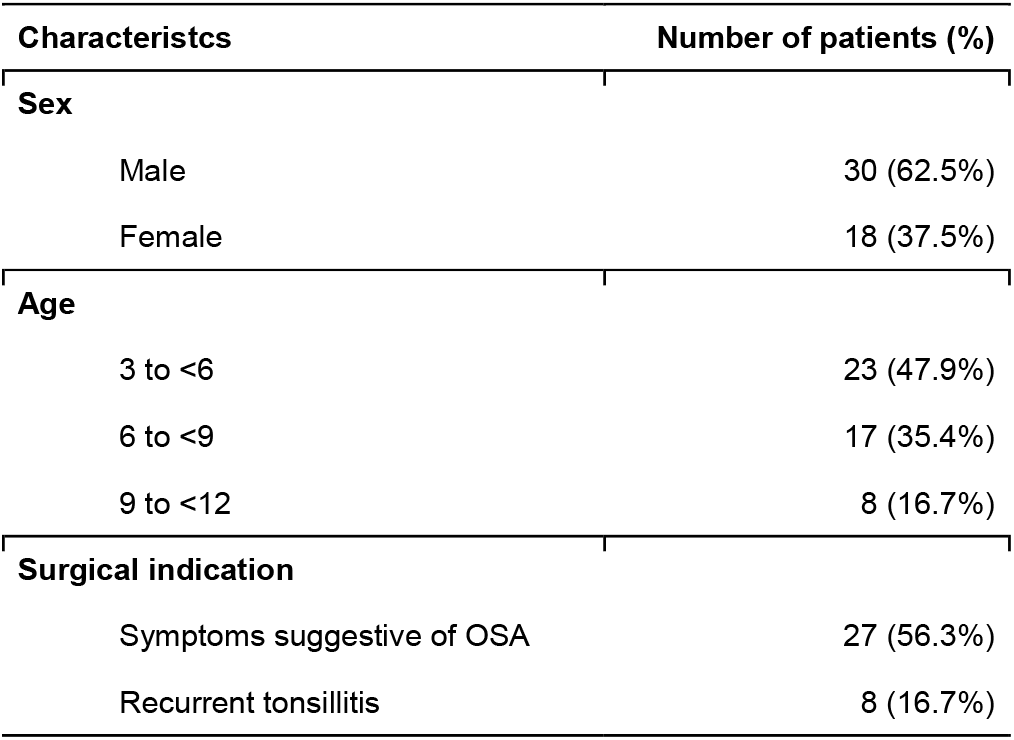
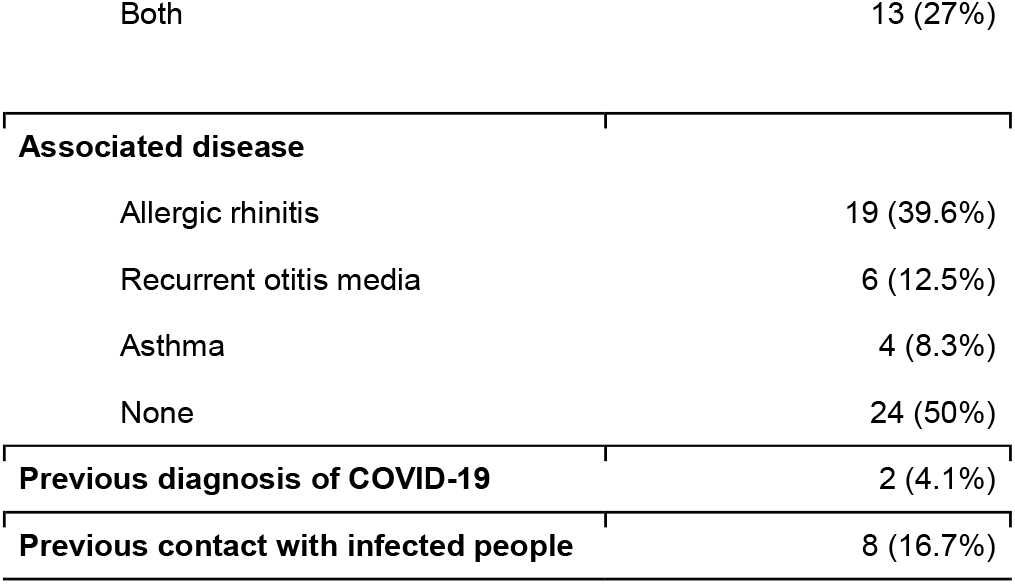
Demographic data from the patients enrolled in this study

### 3.2. Detection of SARS-CoV-2 RNA and antigen

SARS-CoV-2 RNA was detected by RT-PCR in at least one sample from 12 of the 48 patients (25%). In the majority of them, more than one sample was positive (Table 3). The SARS-CoV-2 detection rates were 20% in palatine tonsils, 16% in adenoids, 10% in nasal cytobrushes, and 6% in nasal washes. SARS-CoV-2 viral loads varied widely from 186 to 7114 copies of genome equivalents per μg RNA considering tonsillar tissues, nasal washes and nasal cytobrushes (Figure 1A), and although the median viral load in tonsillar tissues was about two-fold higher than in nasal specimens, there was no significant difference among samples.

**Table 3.** Sample types, distribution of Ct values, serological diagnosis and comorbidity information of patientes with positive SARS-CoV-2.

| Number | Lymphoid tissue/secretions positive | Ct for Gene E | Ct for Gene N2 | Neutralizing antibodies | Serological test (IgG/IgM) | Comorbidities |
| --- | --- | --- | --- | --- | --- | --- |
| 02 | Palatine tonsil | 35,0 | 34,9 | None | None | Allergic rhinitis |
|  | Nasal cytobrush | 36,0 | 35 |  |  |  |
| 04 | Palatine tonsil | 32,6 | 32,2 | None | None | Allergic rhinitis |
|  | Adenoid | 31,1 | 30,7 |  |  |  |
|  | Nasal cytobrush | 35,0 | 34,0 |  |  |  |
| 10 | Adenoid | 32,7 | 30,7 | ND | ND | Others infections |
| 13 | Palatine tonsil | 34,1 | 32,3 | None | None | Asthma |
|  | Adenoid | 32,9 | 32,7 |  |  |  |
| 18 | Palatine tonsil | 28,5 | 25,3 | 1/4 | None | None |
|  | Nasal cytobrush | 31,6 | 25,6 |  |  |  |
| 21 | Nasal wash | 31,2 | 29,7 | 1/32 | None | Allergic rhinitis |
| 26 | Palatine tonsil | 31,2 | 29,1 | None | IgG | None |
|  | Adenoid | 28,8 | 26,5 |  |  |  |
|  | Nasal wash | 31,4 | 28,7 |  |  |  |
| 28 | Palatine tonsil | 31,4 | 28,3 | 1/4 | None | Serous otitis media |
|  | Nasal cytobrush | 31,9 | 27,7 |  |  |  |
| 30 | Palatine tonsil | 28,4 | 27,8 | None | IgG | None |
|  | Adenoid | 32,3 | 31,4 |  |  |  |
|  | Nasal cytobrush | 34,2 | 32,9 |  |  |  |
|  | Nasal wash | 32,6 | 31,6 |  |  |  |
| 41 | Palatine tonsil | 33,0 | 31,0 | 1/32 | IgG | None |
|  | Adenoid | 31,0 | 27,0 |  |  |  |
| 46 | Adenoid | 35,4 | 33,8 | 1/64 | IgG | None |
| 52 | Palatine tonsil | 33,0 | 29,0 | 1/32 | IgG | Allergic rhinitis |

**ND: Not done**

**Figure 1.**
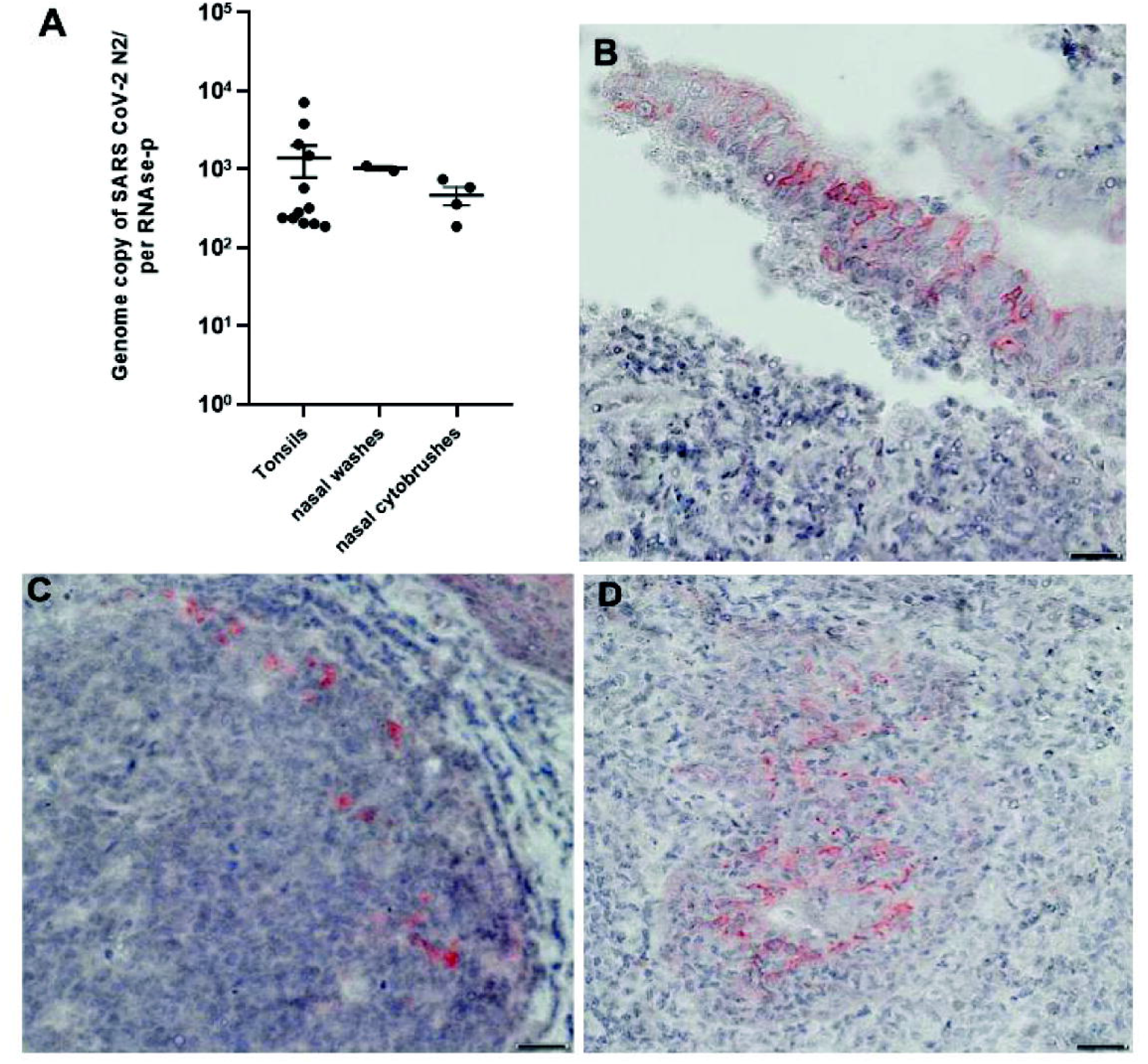
SARS-CoV-2 viral loads and antigen detection. A) Quantification of SARS CoV-2 RNA genome copies in palatine tonsil and adenoid tissues, nasal washes and nasal cytobrushes from patients with chronic adenotonsillar disease (+SD). B). Representative section of an adenoid positive for SARS-CoV-2 NP protein in pseudo-stratified ciliated epithelium, showing SARS-CoV-2-infected cells with red positive signal. C). Representative section of a palatine tonsil positive for SARS-CoV-2 NP protein in a lymphoid follicle. D) Representative section of a palatine tonsil positive for SARS-CoV-2 NP protein in the interfollicular area. Scale bar:100 μm

SARS-CoV-2-positive samples were detected in patients without previous history (personal or in household member) of exposure to SARS-CoV-2, whereas negative samples were found in patients who had been exposed to confirmed COVID-19 cases in the family one to five months before surgery. Five of 12 SARS-CoV-2-positive children had no comorbidities, and the most frequent comorbidity was allergic rhinitis (4 of 12) (Table 3).

Immunohistochemistry was done on all tissue sections from SARS-CoV-2-positive tonsils, and two thirds of them were positive for SARS-CoV-2 antigen, which was detected in tonsillar epithelia and also in scattered cells in the lymphoid compartment, including lymphoid follicles and extrafollicular areas, in both tonsil types (Figure 1B, 1C, and 1D).

### 3.2. Expression of ACE2 and TMPRSS2 in tonsils

Immunofluorescence revealed that the expressions of the main SARS□CoV□2 receptor (ACE2) and S-cleaving transmembrane protease (TMPRSS2) in tonsillar tissue sections was significantly more intense in tissues positive for SARS-CoV-2 as compared to negative ones (Figure 2A). The same areas of enhanced expression were also positive for the SARS-CoV-2 spike protein. The mean differences in fluorescence intensity between SARS-CoV-2-infected and non-infected patients was 4815±840.8 for ACE2 and 9471±847.5 for TMPRSS2 (p<0.0001) (Figure 2B).

**Figure 2.**
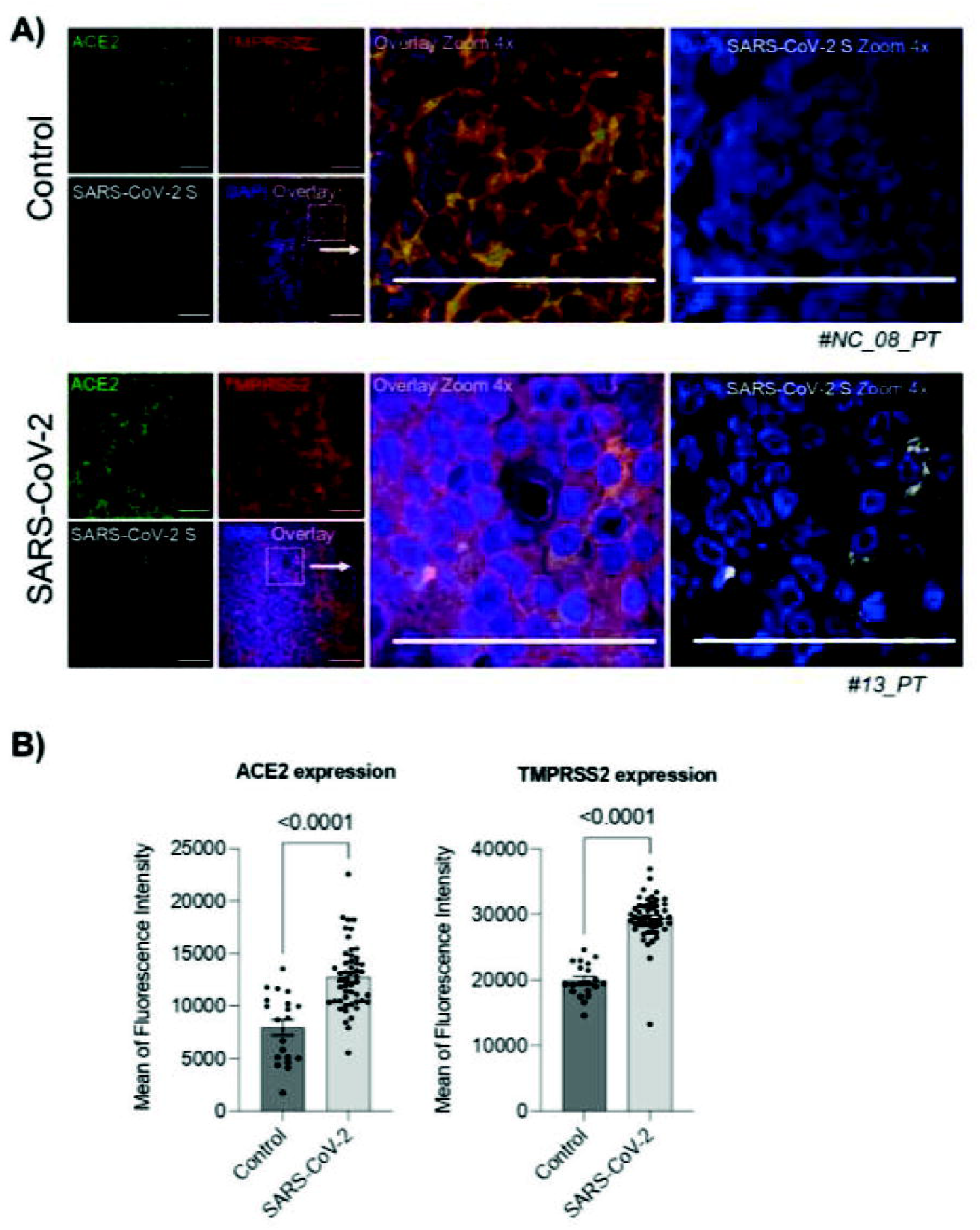
Expression of ACE2 and TMPRSS2 in tonsils. A). Palatine tonsil staining for ACE2, TMPRSS2 and the S protein of SARS-CoV-2. B). Mean fluorescence intensity of ACE2 and TMPRSS2 in SARS-CoV-2-infected tonsils and controls.

### 3.3. Cells infected by SARS-CoV-2 in tonsillar tissues

Gates analises, in Figure 3A, from flow cytometry showed that CD20^+^ B lymphocytes were by far the most frequent cell type, averaging 24.21% and 24% of all SARS-CoV-2-infected TMNCs, respectively in palatine tonsils and adenoids. CD4^+^ T lymphocytes averaged 12.31% and 10.26% of SARS-CoV-2-positive cells, respectively in palatine tonsils and adenoids, and CD8^+^ T lymphocytes averaged 9.87% and 18.18% of SARS-CoV-2-positive cells, respectively in palatine tonsils and adenoids. Among antigen-presenting cells (APC), CD14+ macrophages represented 4.56% and 10.17% SARS-CoV-2-positive cells, respectively in palatine tonsils and adenoids, while CD123+ dendritic cells were the least abundant cell type, with averages of 2.18% and 4.03% of SARS-CoV-2-positive cells, respectively in palatine tonsils and adenoids. In summary, CD20+ B lymphocytes were the most frequent SARS-CoV-2-infected cell in both tonsillar tissue, followed by CD4+, CD8+, CD14+ and CD123+ in palatine tonsils, and by CD8+, CD4+, CD14+ and CD123+ in adenoids (Figure 3).

**Figure 3.**
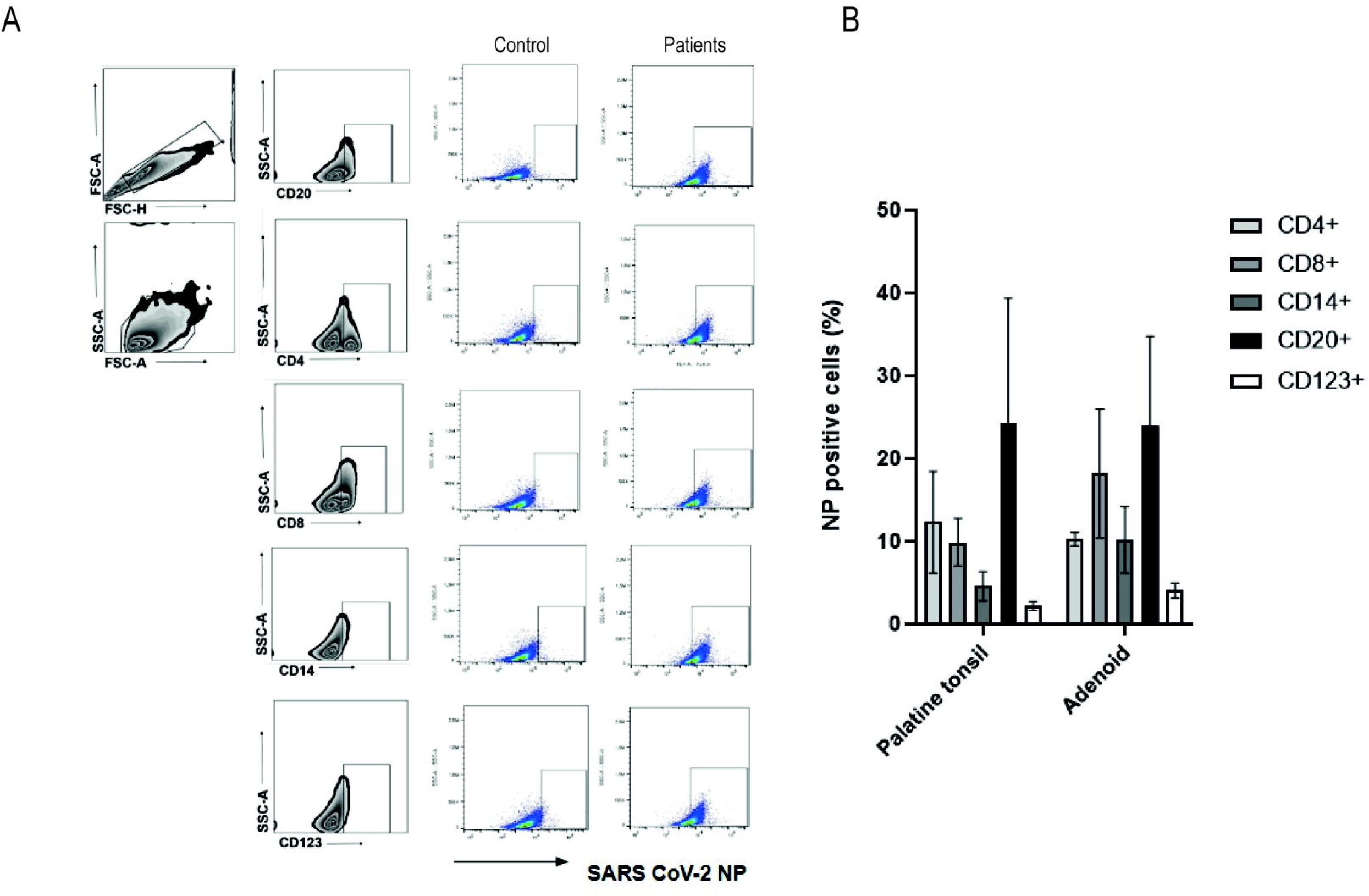
Immune phenotyping of SARS-CoV-2 NP-positive tonsillar cells by flow cytometry. A) A representative gating illustrating the infected population in SARS-CoV-2-negative (Control) and SARS-CoV-2-positive (Patient) tonsils. B) Frequencies of infected immune cells in adenoids and palatine tonsils positive

### 3.4. SARS-CoV-2 RNA and antigen detection in nasal cytobrushes

Cytobrush samples from the olfactory region were positive for SARS-CoV-2 RNA in 5 of the 12 SARS-CoV-2-positive patients, all of whom were also positive in tonsillar tissues. Immunofluorescence revealed SARS-CoV-2-positive cells in cytobrushes from 2 of these 5 patients (Figure 4).

**Figure 4.**
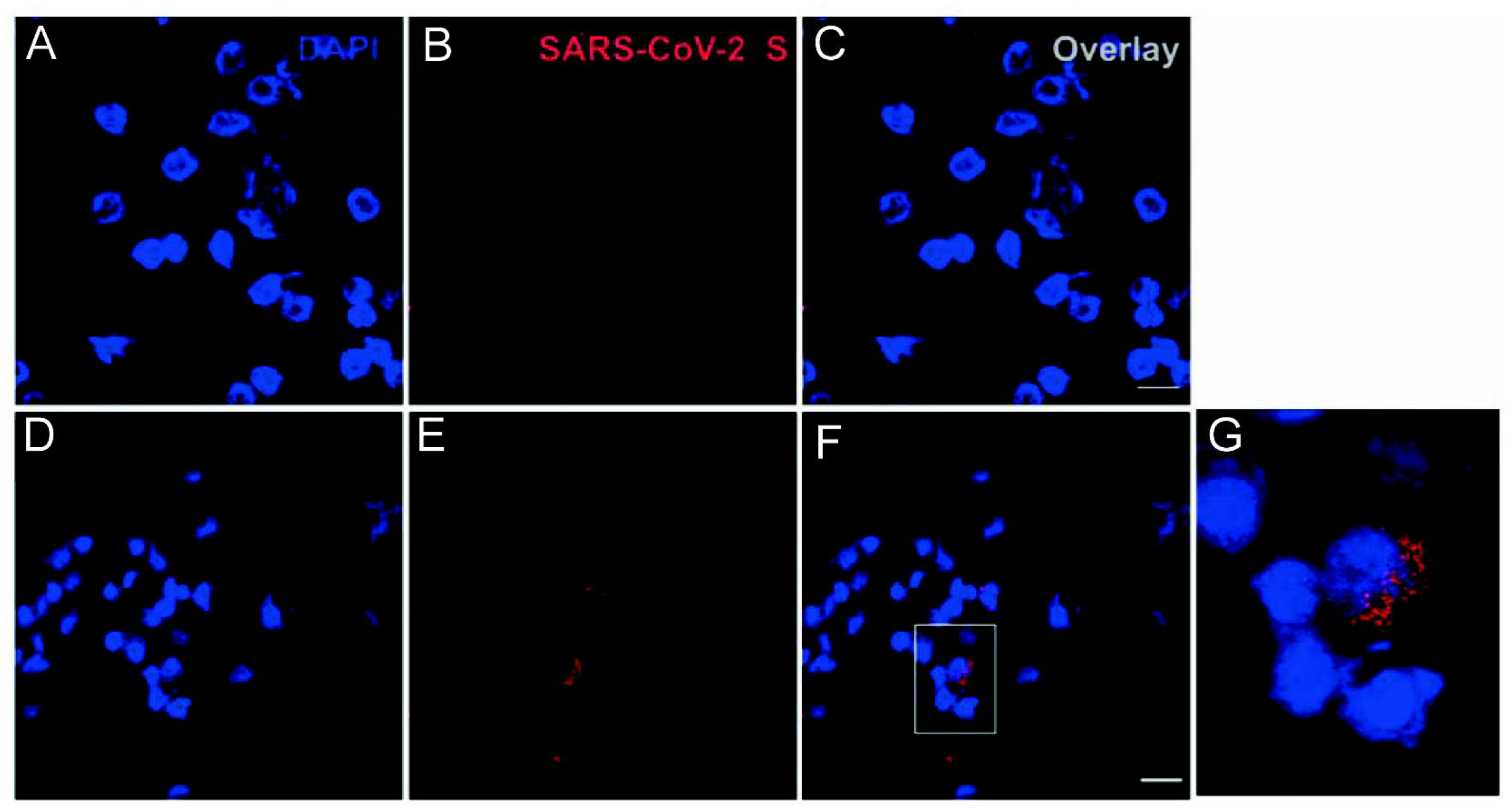
Immunoflurescence for SARS-CoV-2 in cytobrush preparations. Representative fields of cytobrush preparations from SARS-CoV-2-negative (A-C) and SARS-CoV-2-positive children (D-G), showing positivity for NP protein in some cells. The inset on F is enlarged on G. Scale bar: 25 μm.

### 3.5. SARS-CoV-2 serology

Rapid tests for anti-SARS-CoV-2 IgM and IgG antibodies were done in 11 of the 12 SARS-CoV-2-positive patients, whose sera were available, and IgG was detected in 5 patients (45%), while no patient was IgM-positive. This indicates that at least 5 of the 12 SARS-CoV-2-positive patients were not in the acute phase of the infection. Sera from all 5 IgG-positive patients neutralized SARS-CoV-2 in vitro, yet in low titers, as determined by VNT_100_ (Table 3).

### 3.6. SARS-CoV-2 genome sequences

Nanopore sequencing done in 12 samples (9 tonsillar tissues and 3 cytobrushes) from 10 patients yielded SARS-CoV-2 genome sequences in eight children (10 samples). The total length of SARS-CoV-2 sequences varied from 346 to 27,615 nucleotides, with six of them covering less than 40% of the genome length at a 20X depth (Supplementary Table 2). The sequence analysis revealed several Pangolin lineages (Figure 5), all of which circulated in Brazil during the study period.^16^

**Figure 5.**
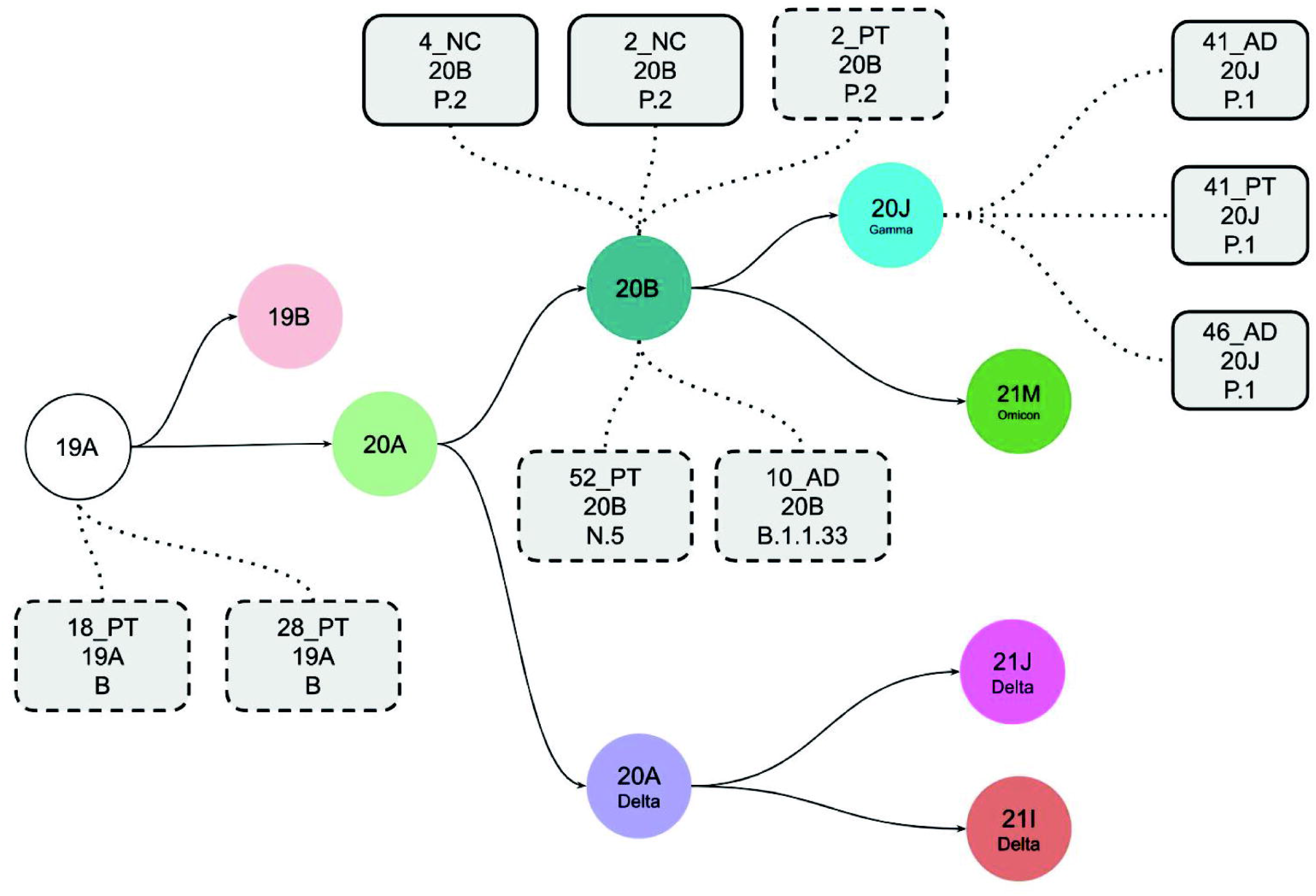
Schematic representation of the evolution of SARS-CoV-2 clades. Each clade is represented by a color circle, and the samples sequenced in the present cohort with the mutational patterns represented by gray rectangles containing the following information, from top to bottom: sample ID, the clade, and the Pangolin lineage assigned by Next Clade. The solid rectangles represent SARS-CoV-2 genomes assembled with higher coverage, and the dashed ones represent those with lower coverage, but carrying enough defining mutations to enable lineage assignment.(AD: adenoid; PT: palatine tonsil; NC: nasal cytobrush).

## 4. DISCUSSION

This study showed that SARS-CoV-2 was detected in upper respiratory tract samples from one quarter of children undergoing tonsillectomy, even in the absence of recent history for COVID-19. This roughly five-fold higher rate than the approximately 5% reported for seasonal coronaviruses in similar cohorts,^4-7^ may result from the sheer intense circulation of SARS-CoV-2 in Brazil in 2021, or from an enhanced propensity of SARS-CoV-2 to infect tonsils, or both.

The time of the patient’s initial exposure to SARS-CoV-2 could not be determined in this cohort, nor was it possible to define a past episode of acute infection for most of the SARS-CoV-2-positive children. Thus, these children can be regarded as asymptomatic SARS-CoV-2 carriers, in agreement with reports that children are more likely than adults to have mild or asymptomatic SARS-CoV-2 infections.^17, 18^ The lower severity of COVID-19 in children can be attributed, at least in part, to some degree of cross-protection afforded by memory T-cell responses to previous infections by endemic coronaviruses.^19^ Also, a more vigorous innate immune response to SARS-CoV-2 in children than in adults could more efficiently contain the agent at the portal of entry, curb the spread to other tissues and reduce illness severity.^20^

SARS-CoV-2 RNA was detected by qRT-PCR in more than one sample from some of the virus-positive children, with viral loads varying from hundreds to thousands of copies per microgram of RNA, suggesting that they may have undergone tonsillectomy at different times post-infection. Nevertheless, the time of initial exposure to SARS-CoV-2 was unknown and most children had no clear symptomatic phase, which hampers the establishment of correlations between duration of infection and viral loads at the time of tonsillectomy. It is important to stress that 5 of the 12 children were IgG-positive, but none was IgM-positive for SARS-CoV-2.

In addition to the detection of SARS-CoV-2 RNA, which could be regarded as some remnant from a past infection, the present study revealed structural viral protein *in situ* in adenoids and palatine tonsils, in both epithelial and lymphomononuclear cells of different lymphoid compartments. This information is novel and provides further evidence for the presence of viral protein synthesis, hence viral activity, in tonsils of children without overt COVID-19. Also, the presence of SARS-CoV-2 protein in cells from the olfactory region in two children who also had virus detected in tonsils, indicates that prolonged SARS-CoV-2 infection is not restricted to tonsillar cells.

SARS-CoV-2 antigen was detected by flow cytometry in the major types of TMNCs, including B and T lymphocytes, macrophages and dendritic cells. Remarkably, this agrees with our previous report that SARS-CoV-2 infects the same range of peripheral blood mononuclear cells (PBMC) from adult COVID-19 patients, inducing apoptosis of infected cells, thus contributing to lymphopenia.^21, 22^ Also, post-mortem studies revealed SARS-CoV-2 infection in human lymphomononuclear cells, with histological alterations in spleens, lymph nodes, and gut-associated lymphoid tissue.^23, 24^ Those findings, along with the present observations in asymptomatic children, suggest that SARS-CoV-2-infected lymphomononuclear cells may intermigrate among secondary lymphoid organs, where intense B lymphocyte maturation and T lymphocyte activation take place.

ACE2 and TMPRSS2 proteins are highly expressed in the upper respiratory tract,^25^ and the present finding of even higher ACE2 and TMPRSS2 expressions in SARS-CoV-2-infected tonsils may suggest that SARS-CoV-2 tonsillar infection promotes increased expression of ACE2 and TMPRSS2. Alternatively, a higher constitutive expression of ACE2 and TMPRSS2, depending on individual variation, could predispose some children to SARS-CoV-2 infection in tonsils.

B lymphocytes comprised roughly one quarter of SARS-CoV-2-infected TMNCs in both types of tonsils, which is not surprising, considering that they are the most abundant cells in secondary lymphoid organs. Of note, B lymphocytes were also the most frequently infected cells in PBMCs from acute COVID-19 adult patients.^21^

The median frequencies of CD8+T lymphocytes in SARS-CoV-2-infected TMNCs were respectively 10% and 18% in palatine tonsils and adenoids, consistent with the rates found in PBMCs from acute COVID-19 patients.^21^ The infection of CD8+T lymphocytes by viruses is surprising, considering that these are the very cells that perform cytotoxicity of virus-infected cells, and thus are central in the combat of viral infections. We have previously reported silent infection of tonsillar CD8+T lymphocytes also by influenza A virus,^26^ suggesting that infection of these cells by viruses may have been previously overlooked. It has been suggested that SARS-CoV-2 persistence may be associated with a virus-specific CD8+T cell response,^27^ which was not assessed in the present study.

SARS-CoV-2 was also detected in tonsillar CD14^+^ monocytes and CD123^+^ dendritic cells, which play important roles as components of the innate immune response. It is presently unknown whether SARS-CoV-2 in APCs results from virus antigen internalization, or to their permissiveness to SARS-CoV-2 replication, or both. Considering that monocytes, macrophages and dendritic cells are infected by SARS-CoV-2, and that infection of human monocytes triggers inflammasomes,^31^ the infection of such cells in tonsils perhaps enhances inflammation in an already chronically inflamed tissue.^28, 29^

At present, it is unknown whether SARS-CoV-2 infection of lymphocytes and APCs is detrimental to their function in secondary lymphoid tissues. Moreover, the antigenic specificities of infected lymphocytes in secondary lymphoid organs, and whether some of these cells are naïve, memory, or innate cells, is also presently unknown.

In the present study, SARS-CoV-2 genome was detected by qRT-PCR in nasal washes from 3 of the 12 SARS-CoV-2-positive children, in the absence of COVID-19 symptoms. This agrees with reports that up to 50% of children under 11 years with SARS-CoV-2 infection may be asymptomatic.^30^ Therefore, the findings suggest that tonsils and olfactory epithelium may be sources of SARS-CoV-2 shedding in nasal washes of asymptomatic children, who may be sources of virus transmission for the community.^31^

Variable loads of SARS-CoV-2 RNA are shed in nasopharyngeal secretions and saliva from COVID-19 patients, including asymptomatic ones.^32^ In the present cohort, the median viral loads were not significantly different among different sampling sites.

It is noteworthy that two of twelve SARS-CoV-2-positive children had a previous laboratory-confirmed SARS-CoV-2 infection, dating back three and five months prior to tonsillectomy, indicating that they had prolonged or persisting SARS-CoV-2 infection. In further support of SARS-CoV-2 prolonged or persisting infection in tonsils, rather than reinfection, all five children who were seropositive for antibodies to SARS-CoV-2 at the time of tonsillectomy in the present cohort lacked IgM antibodies. However, since we had no access to backup respiratory samples collected at the time of acute infection, it was not possible to ascertain whether the virus detected at the time of tonsillectomy of those two children were of the same strain causing the acute infection, or a reinfecting new one.

Several SARS-CoV-2 proteins alter components of the cell transcriptome, proteome, ubiquitinome, and phosphoproteome, to evade host defenses and be able to persist in low-grade infection profile.^33^

Importantly, genome sequencing revealed SARS-CoV-2 of several Pangolin lineages in human tonsils, suggesting that tropism for tonsillar cells is not specific to certain lineages. Whole SARS-CoV-2 genome sequences were not obtained from the infected tissues, which is understandable, considering that the tissue samples may have undergone partial autolysis with viral RNA degradation. In addition, the heterogeneity in the intra-tissular distribution of SARS-CoV-2 RNA among different regions of the tonsils, which were randomly split for the different assays, may have also contributed to that. Nevertheless, the available coverage and sequence depth attained in 10 samples from 8 patients enabled the safe calling of Pangolin lineages.

To the best of our knowledge, this is the first study to identify sites of infection and host cell types of SARS-CoV-2, in tissues where the agent may remain for prolonged times, clinically silent, in the upper respiratory tract of children. In addition to SARS-CoV-2 RNA, viral protein was also detected in tonsils, adenoids, and nasal epithelial cells, an evidence of translational activity in situ. Besides epithelial cells, all major types of lymphomononuclear cells host SARS-CoV-2, which may contribute to the maintenance of SARS-CoV-2 in lymphoid tissues of the upper respiratory tract, with still unknown potential immunoinflammatory consequences. These findings underpin the potential role of hypertrophic tonsils as sites of SARS-CoV-2 infection in children, for an undetermined prolonged time. Such smoldering SARS-CoV-2 infection might involve continuous low-level production of viral proteins and cell-to-cell transmission, which circumvent immune surveillance and subvert sterilizing immunity by low virus replication and possibly antigenic variation.

## Data Availability

All data produced in the present study are available upon reasonable request to the authors.

## Acknowledgments

We are indebted to Prof. José Luiz Proenca-Modena (UNICAMP) and Prof. Fabrício Campos (UFRGS) for helpful discussions about the Nanopore sequencing protocol.

---

**Supplementary table 1.**
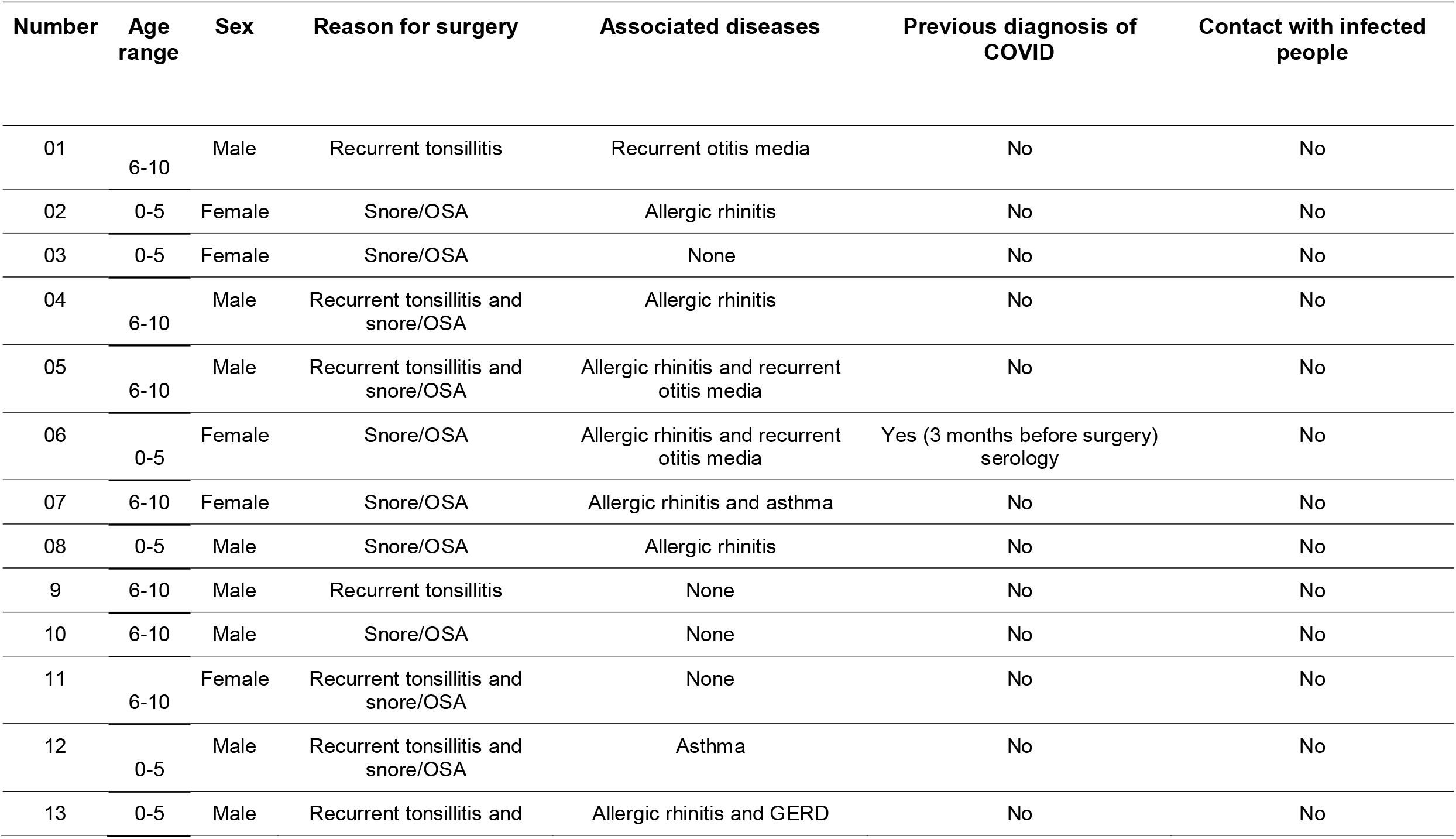
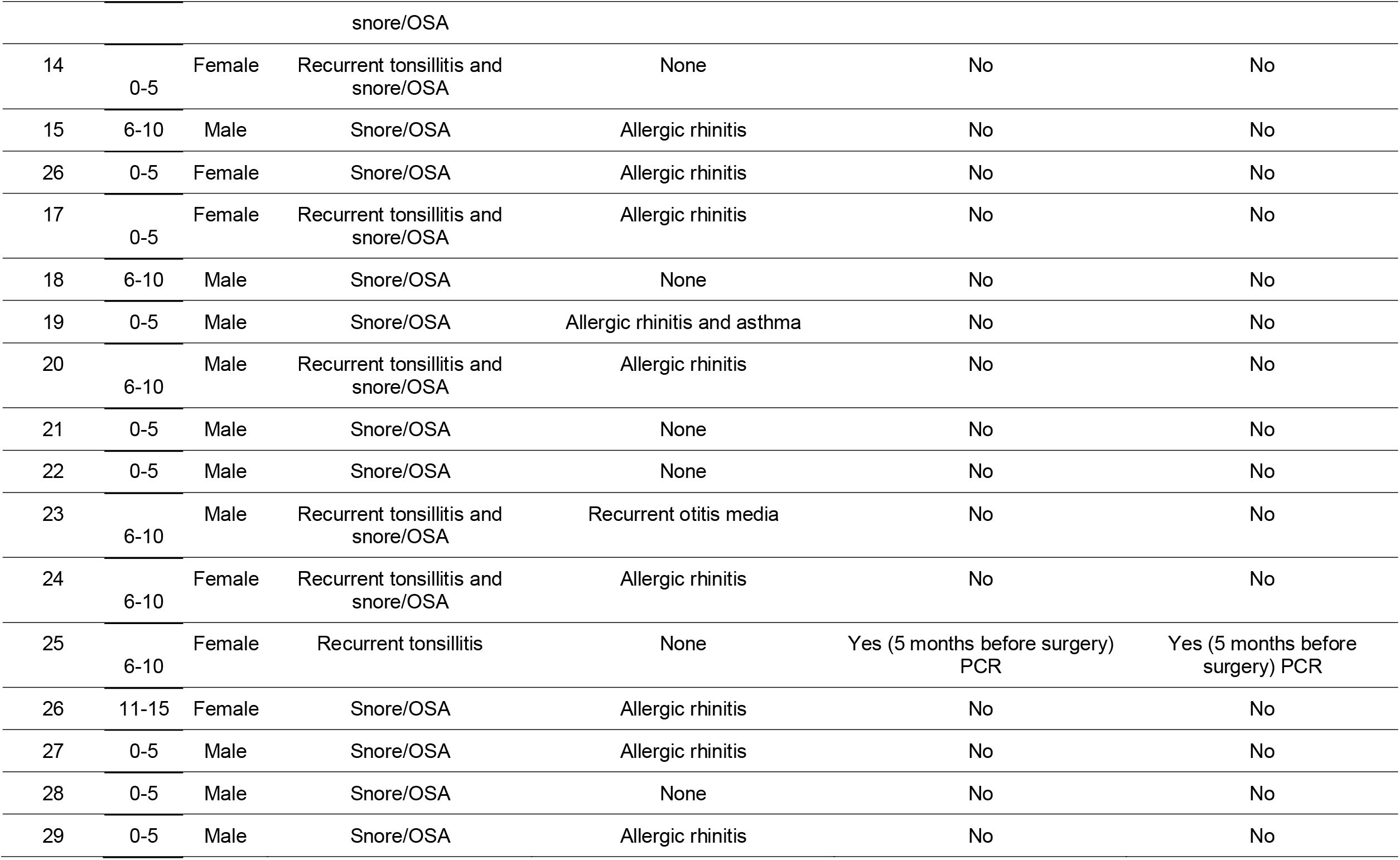
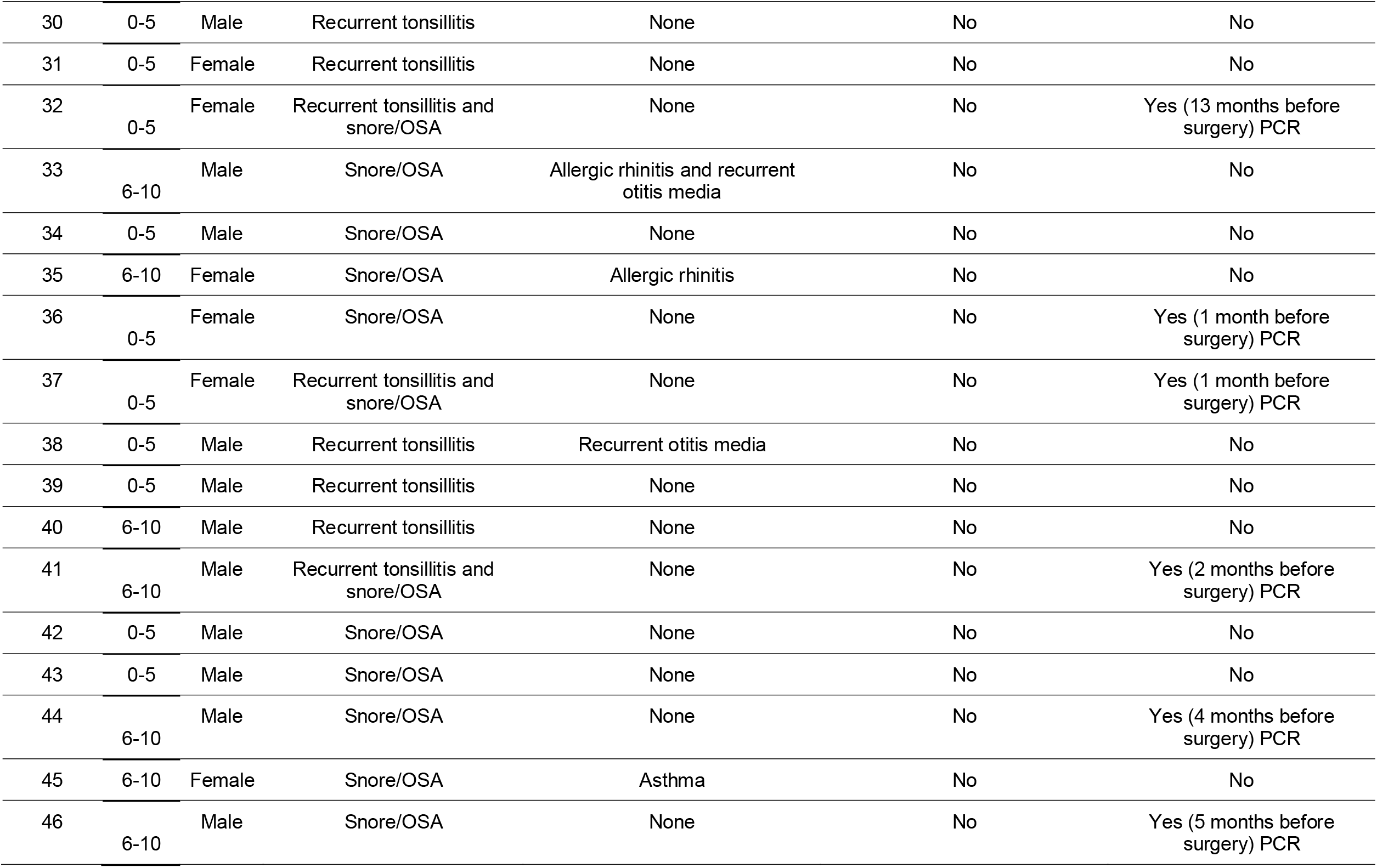
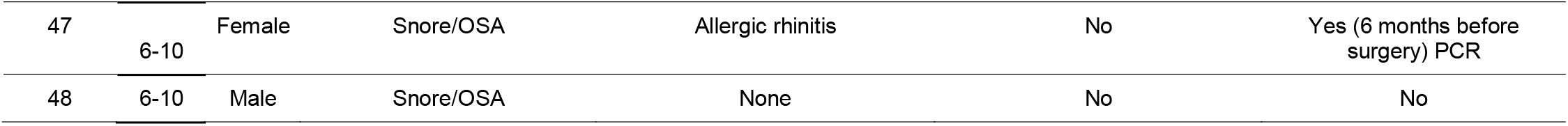
Demographic characteristics and surgery indications for each patient.

| Number | Age range | Sex | Reason for surgery | Associated diseases | Previous diagnosis of COVID | Contact with infected people |
| --- | --- | --- | --- | --- | --- | --- |
| 01 | 6-10 | Male | Recurrent tonsillitis | Recurrent otitis media | No | No |
| 02 | 0-5 | Female | Snore/OSA | Allergic rhinitis | No | No |
| 03 | 0-5 | Female | Snore/OSA | None | No | No |
| 04 | 6-10 | Male | Recurrent tonsillitis and snore/OSA | Allergic rhinitis | No | No |
| 05 | 6-10 | Male | Recurrent tonsillitis and snore/OSA | Allergic rhinitis and recurrent otitis media | No | No |
| 06 | 0-5 | Female | Snore/OSA | Allergic rhinitis and recurrent otitis media | Yes (3 months before surgery) serology | No |
| 07 | 6-10 | Female | Snore/OSA | Allergic rhinitis and asthma | No | No |
| 08 | 0-5 | Male | Snore/OSA | Allergic rhinitis | No | No |
| 9 | 6-10 | Male | Recurrent tonsillitis | None | No | No |
| 10 | 6-10 | Male | Snore/OSA | None | No | No |
| 11 | 6-10 | Female | Recurrent tonsillitis and snore/OSA | None | No | No |
| 12 | 0-5 | Male | Recurrent tonsillitis and snore/OSA | Asthma | No | No |
| 13 | 0-5 | Male | Recurrent tonsillitis and | Allergic rhinitis and GERD | No | No |

|  |  |  | snore/OSA |  |  |  |
| --- | --- | --- | --- | --- | --- | --- |
| 14 | 0-5 | Female | Recurrent tonsillitis and snore/OSA | None | No | No |
| 15 | 6-10 | Male | Snore/OSA | Allergic rhinitis | No | No |
| 26 | 0-5 | Female | Snore/OSA | Allergic rhinitis | No | No |
| 17 | 0-5 | Female | Recurrent tonsillitis and snore/OSA | Allergic rhinitis | No | No |
| 18 | 6-10 | Male | Snore/OSA | None | No | No |
| 19 | 0-5 | Male | Snore/OSA | Allergic rhinitis and asthma | No | No |
| 20 | 6-10 | Male | Recurrent tonsillitis and snore/OSA | Allergic rhinitis | No | No |
| 21 | 0-5 | Male | Snore/OSA | None | No | No |
| 22 | 0-5 | Male | Snore/OSA | None | No | No |
| 23 | 6-10 | Male | Recurrent tonsillitis and snore/OSA | Recurrent otitis media | No | No |
| 24 | 6-10 | Female | Recurrent tonsillitis and snore/OSA | Allergic rhinitis | No | No |
| 25 | 6-10 | Female | Recurrent tonsillitis | None | Yes (5 months before surgery) PCR | Yes (5 months before surgery) PCR |
| 26 | 11-15 | Female | Snore/OSA | Allergic rhinitis | No | No |
| 27 | 0-5 | Male | Snore/OSA | Allergic rhinitis | No | No |
| 28 | 0-5 | Male | Snore/OSA | None | No | No |
| 29 | 0-5 | Male | Snore/OSA | Allergic rhinitis | No | No |
| 30 | 0-5 | Male | Recurrent tonsillitis | None | No | No |
| 31 | 0-5 | Female | Recurrent tonsillitis | None | No | No |
| 32 | 0-5 | Female | Recurrent tonsillitis and snore/OSA | None | No | Yes (13 months before surgery) PCR |
| 33 | 6-10 | Male | Snore/OSA | Allergic rhinitis and recurrent otitis media | No | No |
| 34 | 0-5 | Male | Snore/OSA | None | No | No |
| 35 | 6-10 | Female | Snore/OSA | Allergic rhinitis | No | No |
| 36 | 0-5 | Female | Snore/OSA | None | No | Yes (1 month before surgery) PCR |
| 37 | 0-5 | Female | Recurrent tonsillitis and snore/OSA | None | No | Yes (1 month before surgery) PCR |
| 38 | 0-5 | Male | Recurrent tonsillitis | Recurrent otitis media | No | No |
| 39 | 0-5 | Male | Recurrent tonsillitis | None | No | No |
| 40 | 6-10 | Male | Recurrent tonsillitis | None | No | No |
| 41 | 6-10 | Male | Recurrent tonsillitis and snore/OSA | None | No | Yes (2 months before surgery) PCR |
| 42 | 0-5 | Male | Snore/OSA | None | No | No |
| 43 | 0-5 | Male | Snore/OSA | None | No | No |
| 44 | 6-10 | Male | Snore/OSA | None | No | Yes (4 months before surgery) PCR |
| 45 | 6-10 | Female | Snore/OSA | Asthma | No | No |
| 46 | 6-10 | Male | Snore/OSA | None | No | Yes (5 months before surgery) PCR |
| 47 | 6-10 | Female | Snore/OSA | Allergic rhinitis | No | Yes (6 months before surgery) PCR |
| 48 | 6-10 | Male | Snore/OSA | None | No | No |

**Supplementary table 2.** Total Assembled genomes of Sars-CoV-2 obtained by MinIon Sequencing with total mutation numbers compared with Reference genome (MN908947.3).

| Number | Sample | Reads | Total reads (HAC) | Total Mapped Reads (HAC) | Coverage (100X) | Coverage (>20X) | Sequence Total Length | Mapped Percentage (MN908947.3) | Nucleotide Subs | Aminoacid Subs | GAPS | Clade (nextclade) | Lineage (Pangolin) | BioSample (NCBI) |
| --- | --- | --- | --- | --- | --- | --- | --- | --- | --- | --- | --- | --- | --- | --- |
| 02 | palatine tonsils | 173077 | 180435 | 19586 | 16,3 | 20 | 5522 | 10,85 | 6 | 4 | 0 | 20B | P.2 | SAMN30649075 |
| 02 | nasal cytobrush | 179706 | 211176 | 101854 | 67,9 | 80,7 | 22256 | 48,23 | 18 | 9 | 0 | 20B | P.2 | SAMN30649076 |
| 04 | nasal cytobrush | 185466 | 232057 | 204188 | 89,1 | 95,3 | 27615 | 87,99 | 20 | 11 | 0 | 20B | P.2 | SAMN30649077 |
| 10 | adenoid | 8942 | 7103 | 2371 | 4,2 | 6,8 | 1740 | 33,38 | 1 | 1 | 0 | 20B | B.1.1.33 | SAMN30649069 |
| 18 | palatine tonsils | 172135 | 193224 | 7879 | 1,3 | 2,6 | 653 | 4,08 | 0 | 0 | 0 | 19A | B | SAMN30649072 |
| 20 | palatine tonsils | 40846 | 13748 | 19 | 0 | 0 | - | 0,14 | - | - | - | failure* | - | - |
| 28 | palatine tonsils | 230408 | 246127 | 91963 | 20,1 | 20,4 | 5220 | 37,36 | 1 | 0 | 0 | 19A | B | SAMN30649070 |
| 28 | nasal cytobrush | 167552 | 84898 | 5 | 0 | 0 | 0 | 0,01 | - | - | - | failure* | - | - |
| 30 | adenoid | 92205 | 108205 | 21 | 0 | 0 | - | 0,02 | - | - | - | failure* | - | - |
| 41 | palatine tonsils | 67270 | 80365 | 55040 | 30,4 | 43,3 | 11215 | 68,49 | 20 | 12 | 0 | 20J (Gamma, V3) | P.1 | SAMN30649079 |
| 41 | adenoid | 246159 | 265145 | 313783 | 80 | 88 | 25580 | 84,5 | 33 | 18 | 9 | 20J (Gamma, V3) | P.1 | SAMN30649078 |
| 46 | adenoid | 150776 | 191253 | 73641 | 29,1 | 30,9 | 7906 | 38,5 | 9 | 8 | 0 | 20J (Gamma, V3) | P.1 | SAMN30649080 |
| 52 | palatine tonsils | 5492 | 879 | 34 | 0 | 1,2 | 346 | 3,87 | 2 | 0 | - | 20B | N.5 | SAMN30649081 |
| C- | NT-RT | 2039 | - | - | - | - | - | - | - | - | - | - | - | - |
| C+ | Isolate |  |  |  |  |  |  |  | 10 | 6 | 0 | 20B | B.1.1.33 | SAMN32093249 |

**Supplementary Fig. 1.**
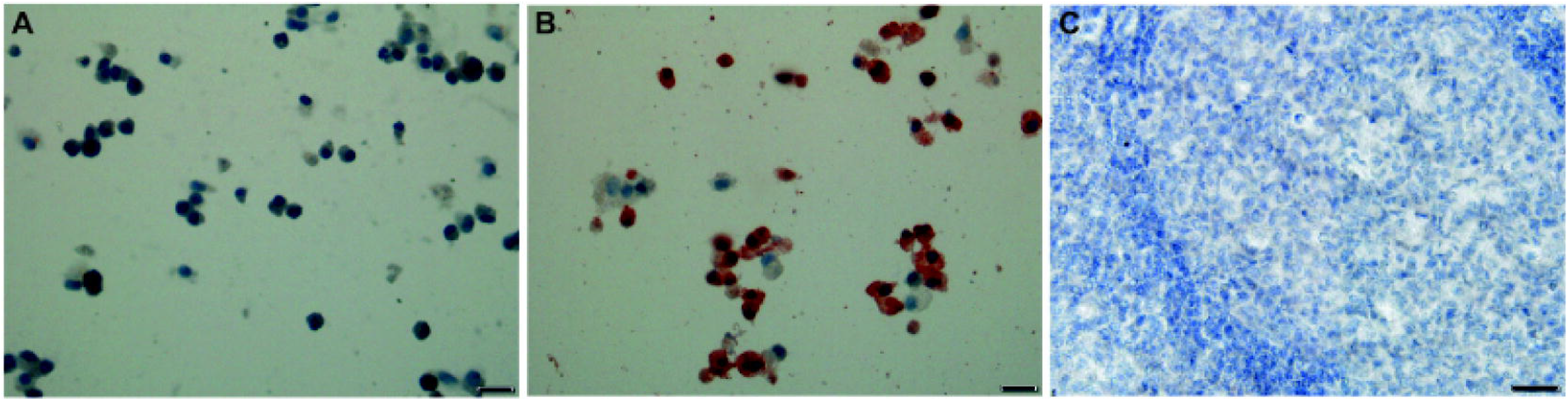
Controls used to standardize immunohistochemistry protocol for SARS-CoV-2 NP protein. A) Non infected Vero CCL-81 cells. B) SARS-Co V-2-infected Vero CCL-81 cells showing abundant reddish signal. C). Representative section of a palatine tonsil negative for SARS-CoV-2, used as negative control, showing absence of signal.

